# Early Pointwise Sensitivity Fluctuation Predicts Glaucoma Progression

**DOI:** 10.64898/2025.12.26.25342592

**Authors:** Jayter Silva Paula, Denny Marcos Garcia, Yuri Ribeiro Carneiro, Micael Oguri dos Reis, Bruna Melchior, Robert N. Weinreb, Linda M. Zangwill, Christopher A. Girkin, Carlos Gustavo De Moraes, Jeffrey M. Liebmann

## Abstract

**Aim:** To assess the relationship between pointwise visual field (VF) sensitivity fluctuation and localised glaucoma progression.

**Methods:** Retrospective observational analysis of prospective cohort data from 399 participants (641 eyes) in the African Descent and Glaucoma Evaluation Study (ADAGES). Glaucoma, glaucoma suspect, and control participants underwent annual examinations including VF testing. VF fluctuation was evaluated using the pointwise standard deviation (SD) of total deviation (TD) residuals during the early 30-month period. Pointwise progression was defined independently at each location as a confirmed sensitivity loss >7 dB. The primary outcome was the association between early fluctuation and subsequent pointwise progression. We additionally evaluated whether the early pointwise rate of change (slope) strengthened this association.

**Results:** Of 33,332 VF points, 5.8% showed progression over 12.2 ± 3.1 years. Progression occurred more frequently in glaucoma (15.6%) than in suspects (1.6%) or controls (0.4%) (p<0.0001). In glaucomatous eyes, progressive points demonstrated greater early fluctuation (median 1.75 dB; IQR 1.52–2.00) than non-progressive points (1.14 dB; IQR 0.97–1.34; p<0.0001) and faster early slopes (–0.65 vs 0.08 dB/year; p<0.0001). In multivariable mixed-effects models, higher early fluctuation (β = 0.40 ± 0.02; p<0.0001) and faster early slopes (β = – 0.40 ± 0.02; p<0.0001), but not baseline TD (p=0.92), were associated with progression. Conclusions: Greater early pointwise VF fluctuation independently predicted future localised progression. The slope analysis mirrored these findings, indicating that early functional variability reflects underlying local instability. These results support early pointwise fluctuation as a predictor of glaucoma progression and a potential endpoint for clinical trials.

## Introduction

Glaucoma is a group of optic neuropathies marked by progressive loss of retinal ganglion cells (RGC) and corresponding visual field (VF) deterioration that may ultimately lead to irreversible blindness.^1^ Standard automated perimetry remains the default method for monitoring glaucomatous functional damage and different criteria have been proposed to detect VF worsening.^2, 3^ Although the psychophysical nature of perimetry results in testing fluctuation, accurate determination of disease severity at diagnosis and close monitoring of VF changes over time are critical for optimizing treatment and follow-up.^4, 5^

Detection of VF progression is challenging due to both short-term and the long-term fluctuations in VF sensitivity, which may mimic or mask true glaucomatous changes.^6,7^ Interestingly, even patients with stable glaucoma presents with some fluctuation in VF sensitivity. However, such fluctuation may be more pronounced in eyes with faster VF decay; which suggests that long-term fluctuation could reflect a latent instability in the visual system.^6,8^ Given this instability is also highest within and adjacent to glaucomatous scotoma it is likely indicative of early RGC loss.^9^

It is therefore important to understand implications of long-term fluctuation, as they may reveal regions of subclinical injury, and predict future glaucoma progression. Thus, while developing methods to address VF sensitivity fluctuation without masking true progression remains important, this variability itself may serve as an early functional surrogate of localized vulnerability to glaucoma progression

Although few studies have focused on early differentiation between population estimates of fluctuation (“noise”) and true VF deterioration (“signal”) to improve clinical decision-making^10–12^, to our knowledge, no study has directly assessed the association between pointwise, sequential VF fluctuation and subsequent glaucoma progression. We propose using the individual eye as its own reference of fluctuation to determine if greater pointwise VF sensitivities variability observed early in follow-up can be used to predict subsequent localized glaucoma progression.

### Subjects and Methods

#### Participants

Clinical data from all participants of the African Descent and Glaucoma Evaluation Study (ADAGES) with follow-up extending through 2019 were used for this study. ADAGES is a multicenter collaboration registered at clinicaltrials.gov under Identifier NCT00221923, which includes the Hamilton Glaucoma Center at the Department of Ophthalmology, University of California-San Diego (UCSD), the Edward S. Harkness Eye Institute at Columbia University Irving Medical Center, and the Department of Ophthalmology at the University of Alabama-Birmingham (UAB). All institutional review boards approved the study, adhering to the Declaration of Helsinki and the Health Insurance Portability and Accountability Act. Informed consent was obtained from all participants.

#### Study Design

ADAGES is an observational, prospective cohort study designed to elucidate factors contributing to disparities in the onset and progression rates of glaucoma between African (AD) and European Descent (ED) patients, whether confirmed or suspected of glaucoma. Treatment approaches were tailored individually by each attending physician. The ocular examinations conducted in ADAGES have been detailed elsewhere.^13^ Participants underwent yearly ophthalmologic evaluations including medical history updates, best-corrected visual acuity checks, slit-lamp biomicroscopy, intraocular pressure (IOP) measurements via Goldmann tonometry, dilated funduscopic exams, pachymetry, and stereoscopic optic disc photography. Standard automated perimetry tests using the 24-2 Swedish interactive threshold algorithm standard (Carl Zeiss Meditec, Inc., Dublin, California, USA) and optic coherence tomography imaging were conducted every six months.

#### Inclusion and Exclusion Criteria

All participants possessed a best-corrected visual acuity ≥ 20/40 and a refractive error within < 5.0 diopters spherical and < 3.0 diopters cylindrical. The inclusion of both eyes was standard unless only one eye fulfilled the criteria.

Exclusion criteria encompassed any ocular or systemic conditions likely to affect the optic nerve or VF results, with ADAGES-defined unreliable VFs, or individuals with fewer than seven VF tests or less than three years of follow-up. Beyond the ADAGES design, stricter VF reliability conditions were used here, defined as > 20% false positives, >33% false negatives, or > 20% fixation losses.

In this study, patients with manifest primary open-angle glaucoma (POAG group) were identified by the presence of eyes with open angles on gonioscopy, glaucomatous optic disc neuropathy based on standard photograph grading protocols of the Imaging Data Evaluation and Analysis (IDEA) reading center, and at least three consecutive abnormal standard automated perimetry tests identified by the Visual Field Assessment Center (VISFaCT) reading center. An abnormal 24-2 VF was defined by a pattern standard deviation P < 5% or a Glaucoma Hemifield Test result “outside normal limits.” Glaucomatous optic neuropathy was identified based on prominent excavation, thinning or notching of the neuroretinal rim, localized or diffuse retinal nerve fiber layer defects, or vertical cup-to-disc ratio asymmetry > 0.2 between eyes (not attributable to disc size differences). These evaluations were made from stereophotographs graded by two independent graders with disagreements resolved by consensus or adjudication.

Eyes presenting IOP levels of 22 mmHg or higher measured on the baseline visits or previously documented and those with sterephotograph based glaucomatous optic neuropathy and normal VF were categorized as glaucoma suspects (suspect group). The control group (assigned as healthy) included eyes with IOP levels lower than 22 mmHg, normal VF, and sterephotograph based optic disc appearance.

#### Visual Field Fluctuation and Progression Assessment

For testing the association between VF sensitivity fluctuation and progression, all 52 values of the total deviation (TD) plot of each 24-2 VF stimulus location points were evaluated. The average of each TD value from the first two tests defined the *pointwise baseline sensitivity. Pointwise sensitivity fluctuation* was analyzed in the first 30 months of follow-up, hereafter called the run-in period. For each VF location point, as from the baseline sensitivity, the standard deviation (SD) of residuals from the linear regression of the TD values based on four or more sequential VF tests of the run-in period was calculated. Potential outliers were excluded using an individual cut-off obtained by calculation of the leverage points.^14^ The result (SD) obtained from each VF location during the run-in period was considered as the eye-specific “fluctuation”, and the median fluctuation for each VF location in the run-in period from all eyes were determined (Figure 1).

**Figure 1.**
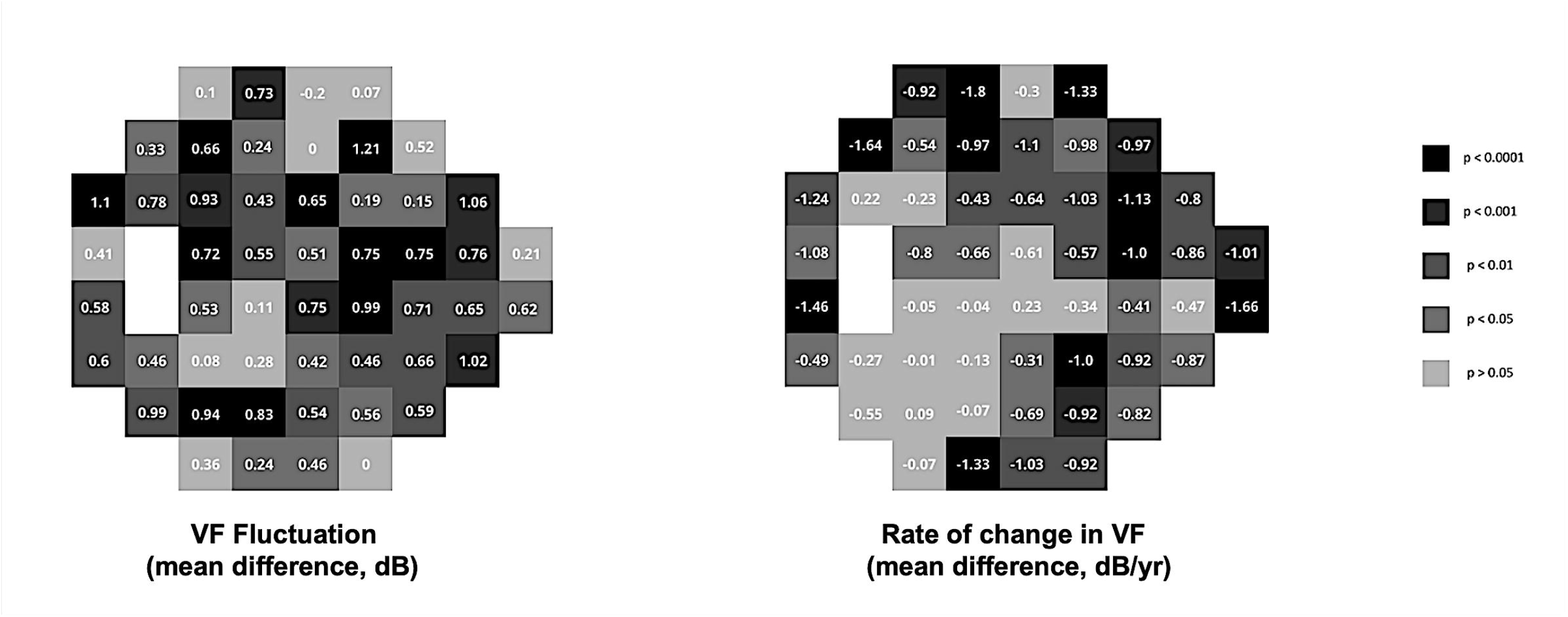
Examples of two 24-2 visual field location points (location N3S9 on the left; location T3S15 on the right) presenting progressive total deviation values worsening in a glaucoma eye over the follow-up period. The total deviation decreases (in dB) that reached the pointwise progression event definition are presented (double-arrows). The slope curves (continuous line through the first black circles) indicate the rate of change in VF (dB/yr) and the insets display the fluctuation (dB) used for calculating the standard deviation of the residuals, up to the initial 30 months (run-in period), as used for comparisons.

Progression was defined based on the functional endpoints discussed at the NEI/FDA Glaucoma Clinical Trial Design and Endpoints Symposium^15–17^ and was considered to have occurred when the sensitivity of any VF test point decreased by more than 7dB after the run-in period, with confirmation on a subsequent VF test.^17^ The association between the run-in pointwise VF fluctuation and further progression was evaluated by comparing the medians of individual SD of residuals between the progressive and non-progressive VF points. Additionally, we compared the pointwise rate of VF sensitivity change (slopes, in dB/year) from each of the VF point sensitivities calculated during the run-in period between the progressive and non-progressive locations using similar approach (Figure 1).

#### Statistical Analysis

Descriptive statistics, including the mean, median, standard deviation (SD), median, standard error (SE) and interquartile ranges (IQR), were used to summarize pointwise sensitivity and sensitivity fluctuations during the run-in period. The exclusion of outliers using leverage points to account for distribution irregularities was performed following ordinary least squares (OLS) linear regression using Python language (version 3.9.13) and Statmodels Python library (version 0.13.2). The Studentized residual for each observation (r⍰) was then computed as r⍰ = e⍰ / [S(⍰)√(1 − h⍰⍰)], where e⍰ is the residual for observation i, S(⍰) is the standard error of the regression excluding that observation, and h⍰⍰ is the leverage value. For comparative analyses, the study utilized non-parametric tests to evaluate the association between both pointwise VF fluctuation and rates of sensitivity change during the run-in period and progression beyond the 7 dB threshold. For each VF test point location across all included eyes during follow-up, we quantified the number of eyes exhibiting progression (“progressive events”) or no progression (“non-progressive events”) at that specific location. Comparisons between progressive and non-progressive events were performed with the Mann-Whitney U test to assess differences in the VF fluctuation and rates of sensitivity change. Heat maps/grayscale maps of 24-2 VF locations were designed using Matplotlib Python library (version 3.5.2). A mixed-effects logistic regression analysis with random-intercept model for each eye was used to verify the association between progression at each VF location and the mean value of the two baseline TD, pointwise VF fluctuation and rates of sensitivity change of the respective location during the run-in period. Significance was set as 5% and the statistical program used was JMP Pro SAS software version 17.0 (SAS Institute Inc.)

## Results

A total of 16,946 VF tests of 641 eyes from 399 ADAGES participants were included. The mean ± SD age of the participants at baseline was 58.6 ± 11.1 years, 59.9% were female (239), 54.9% were classified as European (219 patients), and 45.1% as African descent (180 patients). The mean follow-up period was 12.2 ± 3.1 years (range: 7.5 to 23.3 years) over 17.0 ± 5.9 visits (range: 6 to 41 visits).

We included 364 eyes (56.8%) in the suspect group (baseline VF MD median: −0.4 dB; range: 2.8 to −10.1 dB), 201 eyes (31.4%) in the glaucoma group (baseline VF MD median: −2.3 dB; range: 2.1 to −29.2 dB), and 76 eyes (11.9%) were controls (baseline VF MD median: −0.9 dB; range: 1.5 to −6.7 dB). The pointwise analysis of VF progression encompassed 33,332 individual test points (52 test point locations per eye) and revealed that 94.2% (31,385 points) showed no progression, while 5.8% (1,947 points) exhibited significant worsening based on the repeatable 7 dB criterion (Figure 2).

**Figure 2.**
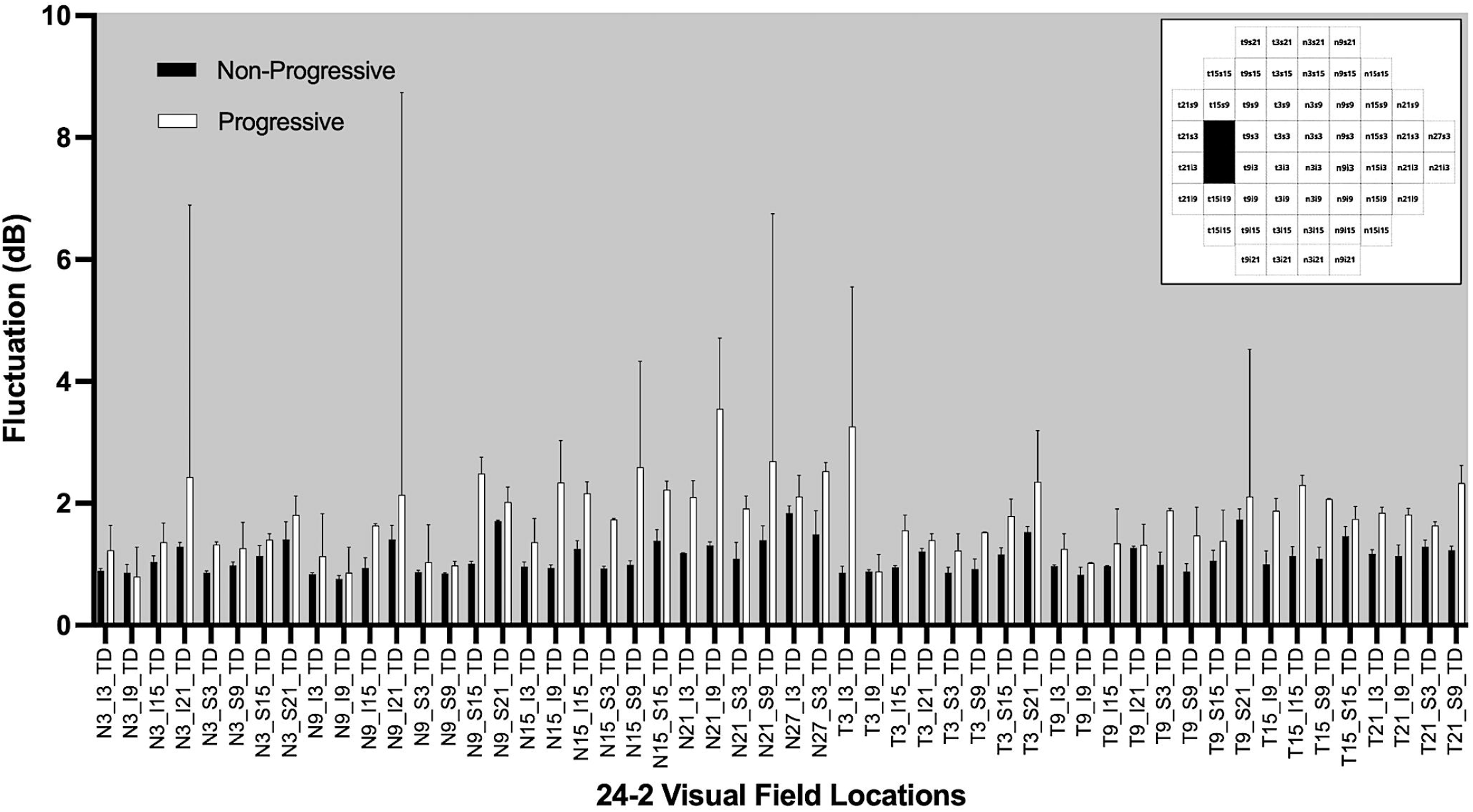
Distribution of the median fluctuation from each of the 24-2 VF test locations from the run-in period, comparing progressive versus non-progressive events. Values are presented as median and 95% confidence interval from all 399 participants (33,332 points tested). *Inset: Illustration of the coordinates’ distribution in 52 test point locations of the 24-2 VF test*.

In the glaucoma group, fewer VF test point locations exhibited progressive events (total: 1,631 events; median: 32 events/location; IQR: 25 to 35 events/location) compared to non-progressive events (total: 8,815 events; median: 169 events/location; IQR: 165 to 176 events/location). In 41 VF test point locations, progressive events presented significantly higher fluctuation than the non-progressive ones (median fluctuation of progressive: 1.75 dB; IQR: 1.52 to 2.00 dB versus non-progressive: 1.14 dB; IQR: 0.97 to 1.34 dB; p<0.0001 – Mann Whitney U test). Two VF locations showed similar median fluctuation in progressive and non-progressive comparison (N3S15: 1.31 dB and N9I21: 1.64 dB), and one location displayed non-significantly higher fluctuation in non-progressive eyes (N3S21: 1.70 versus 1.50 dB; p=0.96) (Table 1, Figures 2 and 3).

**Figure 3.**
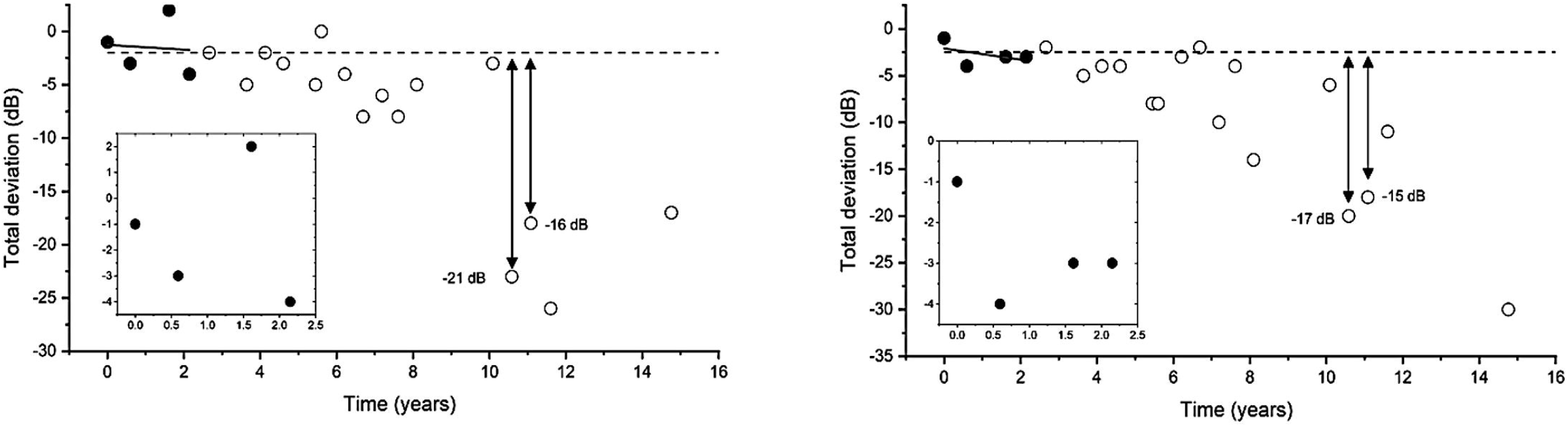
Greyscale maps representing the 24-2 VF test locations depicting different levels of statistical significance, based on p values of the comparison of the medians fluctuation (dB) (left) or run-in rate of change in VF TD (dB/yr) (right) from each 24-2 visual field location, between progression and non-progression events of each point in the glaucoma group. Each square represents a different location and contains the mean difference of standard deviation of TD residuals (fluctuation, left map) or rate of VF sensitivity change (right map) during the run-in period between the progressive and non-progressive events.

In the suspect group, more VF point locations also showed no progression (total: 18,619 events; median: 359 events/location; IQR: 357 to 361 events/location), compared VF to progression (total: 302 events; median: 5 events/location; IQR: 3 to 7 events/location). In 14 VF test point locations (dissimilar locations from the glaucoma group), progressive events presented significantly higher fluctuation compared to non-progressive events (median fluctuation of progressive: 1.71 dB; IQR: 0.98 to 2.23 dB versus non-progressive: 0.99 dB; IQR: 0.87 to 1.21 dB; p<0.0001 – Mann Whitney U test). Eleven VF locations displayed non-significant higher fluctuation in non-progressive (0.87 versus 0.81 dB, p=0.10) (Table 2). Since only a total of 14 events (media: 0 events/location; IQR: 0 to 0 events/location) were deemed progression from 10 VF location points in the control group (versus non-progressive events – median: 76 events/location; IQR: 76 to 76 events/location), comparisons of fluctuation between progressive and non-progressive events were unnecessary (Table 3).

Similarly, a comparison of the pointwise rate of VF TD change during the run-in period in the glaucoma group showed significantly faster rates in progressive events (median sensitivity: – 0.65 dB/year; IQR: –0.92 to –0.25 dB/year) compared to non-progressive events (0.08 dB/year; IQR: 0.00 to 0.18 dB/year; p < 0.0001, Mann–Whitney U test) (Table 4, Figure 3). In the suspect group, significantly faster rates of VF TD change were also seen in progressive VF events (median sensitivity: –0.60 dB/year; IQR: –1.00 to 0.01 dB/year) compared to non-progressive events (median: 0.10 dB/year; IQR: 0.06 to 0.16 dB/year; p < 0.0001, Mann–Whitney U test) (Table 5). Considering the limited number of progressive events in the control group, comparisons of run-in rates of VF TD change between progressive and non-progressive events were not appropriate (Table 6).

Progression was significantly more frequent in the glaucoma group than in glaucoma suspects and controls (15.6% versus 1.6% and 0.4% of points worsened respectively; p <0.0001, chi-square test). The mixed-effects logistic regression showed significant association between each location progression and the run-in period median fluctuations (coefficient ± SE: 0,40 ± 0.02; p<0.0001), and median rate of VF sensitivity change (−0.40 ±0.02; p<0.0001), but not with the mean baseline TD values (0.00 ± 0.01; p=0.92).

## Discussion

In this study, we provide a new insight into the relationship between pointwise VF sensitivity fluctuation and progression, reinforcing the notion that early VF fluctuation may reflect regions of latent functional instability and subthreshold injury in glaucoma and suspect eyes. These findings may also assist in distinguishing true glaucomatous progression and variability due to testing or disease-related fluctuation (“signal-to-noise” ratio).^6, 7^ The use of an eye specific, pointwise analysis approach enabled the identification of specific VF locations that are more likely to show true deterioration, offering an additional tool to conventional trend-based analyses for detecting glaucoma progression.^10, 12^ Taken together, these findings indicate that pointwise VF fluctuation meets key characteristics of a functional biomarker—being measurable, reproducible, and associated with disease risk—and merits further investigation as a future clinical trial endpoint.

The combined analysis of fluctuation magnitude and statistical significance (Figure 3) also supports an assumption of superonasal VF locations might be the most vulnerable sites for early functional instability in glaucoma. These regions demonstrated both larger differences in fluctuation (up to 1.6 dB) and highly significant p-values (p < 0.01) between progressive and non-progressive events. A few discordant findings regarding locations with moderate fluctuation but no statistical significance (e.g., inferotemporal points) suggest potential influences from remaining test variability or individual anatomical differences, warranting further investigation.

Test-retest variability of the VF results in patients with glaucoma has been the subject of numerous studies. Intra-test short-term fluctuation is often related to patient conditions, such as fatigue, anxiety, and fixation issues.^18^ Long-term fluctuation has been associated with additional more complex clinical and biological factors. IOP changes might be correlated to long-term fluctuation, pointing to a potential influence of IOP levels on the optic nerve head perfusion and leading to alterations in VF sensitivities.^19, 20^ Additionally, ocular and systemic vascular dysregulation have been implicated in VF changes.^21^ Systemic factors, such as hypertension, hypotension, and nocturnal blood pressure dips, have also been associated to VF fluctuation.^21, 22^

Moreover, high IOP variability, frequent follow-up exams, prior glaucoma surgery, and faster disease progression were previously reported as potential key predictors of long-term VF fluctuation.^8, 23^ Patients with higher rates of false positives and negatives in VF testing also experienced more fluctuation^8^, highlighting the complex process of discriminating signal to noise in perimetry.

Long-term fluctuation has been shown to be significantly higher in unstable patients compared to those with stable glaucoma^6^ and increased in the later stages of the disease.^24, 25^ Hutchings et al.^7^ further demonstrated that stable glaucoma still exhibited fluctuation, though less than in unstable cases. Nevertheless, results from De Moraes et al.^17^ and Williams et al.^9^ suggest that regions near scotomas may exhibit higher fluctuation sensitivity levels and progression rates compared to normal areas, emphasizing the potential role of VF fluctuation in disease progression.

In our analysis, a significantly stronger association was observed between greater pointwise fluctuation and subsequent VF deterioration among eyes of the glaucoma group compared to suspects and controls. Our findings are consistent with those from previous studies that reported greater long-term fluctuation in patients categorized as nonstable^6^ and suggested that VF worsening was associated with VF fluctuation.^8^ These findings support the notion that VF fluctuation may reflect early functional instability rather than random noise, potentially arising from subclinical reductions in RGC sampling density.^26^ Recognizing such localized sensitivity instability as a manifestation of RGC dysfunction—prior to irreversible loss—may enhance the early identification of regions at risk for glaucomatous progression.^12, 27, 28^ Because early follow-up intervals rarely include enough tests to yield reliable slope estimates, fluctuation would offer a more feasible and informative short-term measure of localized functional behavior. Long-term trend estimation requires extended testing (up to 5-6 years), whereas variability from only 4 tests within 30 months can already reveal meaningful instability. We are aware that pointwise fluctuation might also occur at locations just beginning to deteriorate or undergoing subthreshold progression, but it may nonetheless serve as an early, short functional signature of subsequent worsening.

Our approach to undertaking a more granular analysis of VF fluctuation contrasts with other methods adopted in previous studies, such as those by Mohammadzadeh et al.^11^ and Medeiros et al.^12^ which applied different smoothing methods to adjust for pointwise VF fluctuation during glaucoma progression analysis. Unlike these studies, we applied stringent criteria for VF worsening (7 dB), which might explain the observed high correlation between fluctuation and progression in our cohort.

Although the suspect group showed less progression than the glaucoma group, it also exhibited significant fluctuation linked to VF worsening. These results reinforce a subclinical level of neurodegeneration that is not yet clinically evident as glaucoma deterioration but points to an underlying vulnerability. Prior studies have indicated that such early neurodegenerative changes may be detectable through VF patterns or electrophysiological tests, even before obvious glaucoma manifests. ^6, 29, 30^ These findings could reshape monitoring strategies for glaucoma suspects to utilize early VF fluctuation to enhance the prediction of glaucomatous injury and evaluate subtle neuroenhancement that may occur following IOP reduction.^29–31^ In particular, treatment strategies that reduce this early fluctuation in regions of early RGC injury may be an indicator of neuroenhancement and neuroprotection. These responses may also help refine individualized target pressure selection and identify regions with reversible dysfunction - potentially useful in detecting the effect of both IOP- or non-IOP based therapies. Recent experimental studies have also explored the potential of reversing RGC apoptosis in glaucoma, with some showing that neuroprotective agents or gene therapies may slow or even partially reverse the early degenerative changes in RGC.^31, 32^ Clinically, optical coherence tomography (OCT) and VF studies have demonstrated that lowering IOP not only stabilizes VF loss but can also lead to improvements in retinal nerve fiber layer thickness in some patients, suggesting that early structural changes in glaucoma may be modifiable with adequate IOP control.^22, 33^ These findings underline the complex interplay between IOP reduction and early neuroprotection, reinforcing the need for strategies based on individual variables that display intermittent defects before definitive damage, such as VF fluctuation.

However, our study has limitations. The reliance on retrospective review of data may introduce biases, particularly in the selection of participants and the accuracy of historical management and VF measurements. The exclusion criteria may have eliminated some participants with early or subtle forms of glaucoma, potentially biasing our sample toward more clearly defined cases. Additionally, the definition of VF progression used in this study (a repeatable VF test point sensitivity reduction of 7dB), though robust, might not capture all clinically relevant changes, particularly in early disease stages, where smaller reduction of VF TD may play a more significant role. Notably, our approach of comparing the run-in rate of VF sensitivity change between progressive and non-progressive VF locations, like the Advanced Glaucoma Intervention Study (AGIS) predictive slope-based analysis, strengthened and supported the use of early pointwise VF fluctuation for anticipating disease progression.^34^

Besides, perimetric fluctuation remains a challenge since it may be influenced by psychophysical, technical, and biological factors that are difficult to disentangle. Previous studies from Heijl et al.^18^, Henson et al.^35^, and Zhu et al.^36,37^ have shown that variability depends on sensitivity level and test conditions. However, we speculate that some patients may exhibit fluctuation beyond what psychophysical theory predicts, suggesting a possible component of instability in RGC signaling.

Furthermore, traditional analytical frameworks, using global indices in trend analyses, or unadjusted regression slopes, may cloud localized patterns of instability and failed to remove extreme non-biological outliers. Our residual-based approach along with the limiting approach to extreme outliers intended to mitigate these influences, yet we acknowledge that full separation between biological and methodological variability may be imperfect. Thus, while our results suggest a meaningful biological signal, our conclusions should be accepted within the inherent constraints of VF testing and the multifactorial nature of sensitivity fluctuation.

In conclusion, our study advances the understanding of the relationship between pointwise VF fluctuation and glaucoma progression by highlighting that early fluctuation is not merely random noise but rather a detectable signature of localized susceptibility to functional decline, preceding measurable VF loss.

These findings support the potential of using eye- and point-specific VF analysis as a biomarker for earlier and more precise prediction of glaucoma progression and in the detection of treatment effects on RGC function. While additional studies are needed to validate these findings and refine the prediction models for clinical use, these approaches could ultimately lead to more targeted and effective management strategies for patients at the highest risk of visual deterioration.

## Supporting information

file:///Users/yuriribeiro/Downloads/Table%206.pdf

file:///Users/yuriribeiro/Downloads/Table%205.pdf

file:///Users/yuriribeiro/Downloads/Table%204.pdf

file:///Users/yuriribeiro/Downloads/Table%203.pdf

file:///Users/yuriribeiro/Downloads/Table%202.pdf

file:///Users/yuriribeiro/Downloads/Table%201.pdf

## Data Availability

The data used in this study were obtained from the African Descent and Glaucoma Evaluation Study (ADAGES) database, a multicenter study registered at ClinicalTrials.gov (Identifier: NCT00221923). Restrictions apply to the availability of these data, which were used under authorization for the current study and are not publicly available. Data are, however, available from the ADAGES investigators upon reasonable request and with appropriate institutional approvals.

## References

1. Weinreb RN, Aung T, Medeiros FA. The pathophysiology and treatment of glaucoma: a review. JAMA. 2014;311:1901–1911.

2. Chauhan BC, Malik R, Shuba LM, et al. Practical recommendations for measuring rates of visual field change in glaucoma. Br J Ophthalmol. 2008;92:569–573.

3. Heijl A, Bengtsson B, Chauhan BC, et al. A comparison of visual field progression criteria of 3 major glaucoma trials in early manifest glaucoma trial patients. Ophthalmology. 2008 Sep;115(9):1557–1565.

4. Susanna R Jr, De Moraes CG, et al. Why do people (still) go blind from glaucoma? Transl Vis Sci Technol. 2015;4:1.

5. Chauhan BC, Malik R, Shuba LM, et al. Rates of glaucomatous visual field change in a large clinical population. Invest Ophthalmol Vis Sci. 2014;55:4135–4143.

6. Boeglin RJ, Caprioli J, Zulauf M. Long-term fluctuation of the visual field in glaucoma. Am J Ophthalmol. 1992;113(4):396–400. doi:10.1016/s0002-9394(14)76161-6.

7. Hutchings N, Wild JM, Hussey MK, Flanagan JG, Trope GE. The long-term fluctuation of the visual field in stable glaucoma. Invest Ophthalmol Vis Sci. 2000;41(11):3429–3436.

8. Rabiolo A, Morales E, Kim JH, et al. Predictors of long-term visual field fluctuation in glaucoma patients. Ophthalmology. 2020;127(6):739–747.

9. Haefliger IO, Zobrist T, Flammer J. Fluctuation of the differential light threshold at the border of absolute scotomas. Ophthalmology. 1991;98(10):1529–1532.

10. Gracitelli CPB, Zangwill LM, Diniz-Filho A, et al. Detection of glaucoma progression in individuals of African descent compared with those of European descent. JAMA Ophthalmol. 2018;136(4):329–335.

11. Mohammadzadeh V, Li L, Fei Z, et al. Efficacy of smoothing algorithms to enhance detection of visual field progression in glaucoma. Ophthalmol Sci. 2023;4(2):100423.

12. Medeiros FA, Malek DA, Tseng H, et al. Short-term detection of fast progressors in glaucoma: The Fast Progression Assessment through Clustered Evaluation (Fast-PACE) Study. Ophthalmology. 2024;131(6):645–657.

13. Sample PA, Girkin CA, Zangwill LM, et al. The African Descent and Glaucoma Evaluation Study (ADAGES): design and baseline data. Arch Ophthalmol. 2009;127(9):1136–1145.

14. Thériault R, Ben-Shachar MS, Patil I, et al. Check your outliers! An introduction to identifying statistical outliers in R with easystats. Behav Res Methods. 2024;56(4):4162–4172.

15. Weinreb RN, Kaufman PL. Glaucoma research community and FDA look to the future, II: NEI/FDA Glaucoma Clinical Trial Design and Endpoints Symposium: Measures of structural change and visual function. Invest Ophthalmol Vis Sci. 2011; 52(11), 7842–7851.

16. Levin LA, Sengupta M, Balcer LJ, Kupersmith MJ, Miller NR. Report from the National Eye Institute workshop on neuro-ophthalmic disease clinical trial endpoints: Optic neuropathies. Invest Ophthalmol Vis Sci. 2021; 62(14), 30.

17. De Moraes CG, Lane KJ, Wang X, Liebmann JM. A potential primary endpoint for clinical trials in glaucoma neuroprotection. Sci Rep. 2023;13(1):7098.

18. Heijl A, Lindgren A, Lindgren G. Test-retest variability in glaucomatous visual fields. Am J Ophthalmol. 1989;108(2):130–135. doi:10.1016/0002-9394(89)90006-8

19. Caprioli J, Coleman AL. Intraocular pressure fluctuation: a risk factor for visual field progression at low intraocular pressures in the Advanced Glaucoma Intervention Study. Ophthalmology. 2008;115(7):1123-1129.e3. doi:10.1016/j.ophtha.2007.10.03

20. Nouri-Mahdavi K, Hoffman D, Coleman AL, Liu G, Morales E, Caprioli J. Predictive factors for glaucomatous visual field progression in the advanced glaucoma intervention study. Ophthalmology. 2004;111(9):1627–1635. doi:10.1016/j.ophtha.2004.01.032.

21. Flammer J, Orgul S, Costa VP, et al. The impact of ocular blood flow in glaucoma. Prog Retin Eye Res. 2002;21(4):359–393. doi:10.1016/s1350-9462(02)00008-3.

22. Leske MC, Heijl A, Hyman L, et al. Predictors of long-term progression in the early manifest glaucoma trial. Ophthalmology. 2007;114(11):1965–1972. doi:10.1016/j.ophtha.2007.03.031.

23. Rabiolo A, Morales E, Kim JH, et al. Predictors of long-term visual field fluctuation in glaucoma patients. Ophthalmology. 2020;127(6):739–747. doi:10.1016/j.ophtha.2019.11.021

24. Boeglin RJ, Caprioli J, Zulauf M. Long-term fluctuation of the visual field in glaucoma. Am J Ophthalmol. 1992;113(4):396–400. doi:10.1016/s0002-9394(14)76161-6

25. Fogagnolo P, Sangermani C, Oddone F, et al. Long-term perimetric fluctuation in patients with different stages of glaucoma. Br J Ophthalmol. 2011;95(2):189–193. doi:10.1136/bjo.2010.182758

26. Rabiolo A, Montesano G, Crabb DP, Garway-Heath DF; United Kingdom Glaucoma Treatment Study Investigators. Relationship between intraocular pressure fluctuation and visual field progression rates in the United Kingdom Glaucoma Treatment Study. Ophthalmology. 2024 Aug;131(8):902–913.

27. Chrysostomou V, Hatch RJ, Colgan T, et al. Visuelle Regeneration als Ziel für das Glaukom [Visual recovery as the target for glaucoma]. Ophthalmologe. 2019 Jan;116(1):14–17.

28. Shibeeb O, Chidlow G, Han G, Wood JP, Casson RJ. Effect of subconjunctival glucose on retinal ganglion cell survival in experimental retinal ischaemia and contrast sensitivity in human glaucoma. Clin Exp Ophthalmol. 2016 Jan-Feb;44(1):24–32.

29. Ventura LM, Porciatti V. Pattern electroretinogram in glaucoma. Curr Opin Ophthalmol. 2006;17(2):196–202.

30. Bode SF, Jehle T, Bach M. Pattern electroretinogram in glaucoma suspects: new findings from a longitudinal study. Invest Ophthalmol Vis Sci. 2011;52(7):4300–4306.

31. Boia R, Vecino CE, Santiago AR. Neuroprotective strategies for retinal ganglion cell degeneration: current status and challenges ahead. Int J Mol Sci. 2020;21(7):2262.

32. Rhee J, Shih KC. Use of gene therapy in retinal ganglion cell neuroprotection: current concepts and future directions. Biomolecules. 2021;11(4):581.

33. Medeiros FA, Weinreb RN, Zangwill LM, et al. Long-term intraocular pressure fluctuations and risk of conversion from ocular hypertension to glaucoma. Ophthalmology. 2008;115(6):934–940.

34. Nouri-Mahdavi K, Hoffman D, Gaasterland D, Caprioli J. Prediction of visual field progression in glaucoma. Invest Ophthalmol Vis Sci. 2004;45(12):4346–4351.

35. Henson DB, Chaudry S, Artes PH, et al. Response variability in the visual field: comparison of optic neuritis, glaucoma, ocular hypertension, and normal eyes. Invest Ophthalmol Vis Sci. 2000;41(2):417–21.

36. Zhu H, Russell RA, Saunders LJ, et al. Detecting changes in retinal function: Analysis with Non-Stationary Weibull Error Regression and Spatial enhancement (ANSWERS). PLoS One. 2014;9(1):e85654.

37. Zhu H, Crabb DP, Ho T, et al. More Accurate Modeling of Visual Field Progression in Glaucoma: ANSWERS. Invest Ophthalmol Vis Sci. 2015;56(10):6077–83.

