## Supplementary material for "Early Pointwise Sensitivity Fluctuation Predicts Glaucoma Progression": file:///Users/yuriribeiro/Downloads/Table%206.pdf

**Table 6.** Comparison of the run-in medians rate of VF sensitivity change (dB) of total deviation (TD) from each 24-2 visual field location, considering progressive and non-progressive events in the control group.

| VF location | Non-progressive events | | | | Progressive events | | | |
|---|---|---|---|---|---|---|---|---|
| | N (eyes) | Median | P25 | P75 | N (eyes) | Median | P25 | P75 |
| N3_I3_TD | 76 | 0.17 | -0.34 | 0.86 | 0 | | | |
| N3_I9_TD | 76 | -0.00 | -0.53 | 0.61 | 0 | | | |
| N3_I15_TD | 76 | 0.23 | -0.37 | 0.84 | 0 | | | |
| N3_I21_TD | 75 | -0.05 | -0.81 | 0.77 | 1 | -5.35 | -5.35 | -5.35 |
| N3_S3_TD | 76 | 0.37 | -0.37 | 0.94 | 0 | | | |
| N3_S9_TD | 76 | 0.28 | -0.49 | 0.79 | 0 | | | |
| N3_S15_TD | 76 | 0.19 | -0.55 | 1.20 | 0 | | | |
| N3_S21_TD | 76 | 0.15 | -0.55 | 1.33 | 0 | | | |
| N9_I3_TD | 76 | 0.20 | -0.23 | 0.75 | 0 | | | |
| N9_I9_TD | 76 | 0.11 | -0.39 | 0.72 | 0 | | | |
| N9_I15_TD | 76 | 0.40 | -0.00 | 0.87 | 0 | | | |
| N9_I21_TD | 75 | 0.00 | -1.04 | 1.10 | 1 | -5.30 | -5.30 | -5.30 |
| N9_S3_TD | 76 | 0.07 | -0.23 | 0.99 | 0 | | | |
| N9_S9_TD | 76 | 0.19 | -0.54 | 0.78 | 0 | | | |
| N9_S15_TD | 76 | 0.11 | -0.22 | 1.21 | 0 | | | |
| N9_S21_TD | 74 | 0.68 | -0.53 | 1.67 | 2 | -3.92 | -7.21 | -0.62 |
| N15_I3_TD | 76 | 0.14 | -0.45 | 0.79 | 0 | | | |
| N15_I9_TD | 76 | 0.09 | -0.41 | 0.77 | 0 | | | |
| N15_I15_TD | 76 | 0.15 | -0.44 | 1.24 | 0 | | | |
| N15_S3_TD | 76 | 0.29 | -0.37 | 1.09 | 0 | | | |
| N15_S9_TD | 75 | 0.33 | -0.01 | 0.96 | 1 | -2.89 | -2.89 | -2.89 |
| N15_S15_TD | 76 | 0.25 | -0.59 | 1.13 | 0 | | | |
| N21_I3_TD | 76 | 0.36 | -0.37 | 0.98 | 0 | | | |
| N21_I9_TD | 76 | 0.38 | -0.38 | 1.04 | 0 | | | |
| N21_S3_TD | 76 | 0.49 | -0.25 | 1.36 | 0 | | | |
| N21_S9_TD | 75 | 0.45 | -0.17 | 1.17 | 1 | -5.77 | -5.77 | -5.77 |
| N27_I3_TD | 76 | 0.27 | -0.23 | 1.33 | 0 | | | |
| N27_S3_TD | 74 | 0.76 | -0.43 | 1.70 | 2 | -3.89 | -5.76 | -2.01 |
| T3_I3_TD | 76 | 0.11 | -0.44 | 0.65 | 0 | | | |
| T3_I9_TD | 76 | 0.08 | -0.40 | 0.93 | 0 | | | |
| T3_I15_TD | 76 | 0.08 | -0.52 | 0.70 | 0 | | | |
| T3_I21_TD | 75 | 0.26 | -0.57 | 0.88 | 1 | -0.87 | -0.87 | -0.87 |
| T3_S3_TD | 76 | 0.20 | -0.37 | 0.76 | 0 | | | |
| T3_S9_TD | 76 | 0.40 | -0.22 | 1.01 | 0 | | | |
| T3_S15_TD | 76 | 0.35 | -0.38 | 1.23 | 0 | | | |
| T3_S21_TD | 75 | 0.35 | -0.78 | 1.69 | 1 | -2.10 | -2.10 | -2.10 |
| T9_I3_TD | 76 | 0.11 | -0.44 | 0.67 | 0 | | | |
| T9_I9_TD | 76 | 0.35 | -0.44 | 0.86 | 0 | | | |
| T9_I15_TD | 76 | 0.27 | -0.27 | 0.82 | 0 | | | |
| T9_I21_TD | 76 | 0.15 | -0.97 | 1.02 | 0 | | | |
| T9_S3_TD | 76 | 0.29 | -0.46 | 0.85 | 0 | | | |
| T9_S9_TD | 76 | 0.26 | -0.34 | 0.82 | 0 | | | |
| T9_S15_TD | 76 | 0.21 | -0.41 | 0.96 | 0 | | | |
| T9_S21_TD | 73 | 0.53 | -0.80 | 1.48 | 3 | -3.88 | -4.53 | 1.27 |

| | | | | | | | | |
|---|---|---|---|---|---|---|---|---|
| T15_I9_TD | 76 | 0.04 | -0.62 | 0.74 | 0 | | | |
| T15_I15_TD | 76 | 0.25 | -0.53 | 1.15 | 0 | | | |
| T15_S9_TD | 76 | 0.33 | -0.48 | 1.13 | 0 | | | |
| T15_S15_TD | 76 | 0.47 | -0.52 | 1.21 | 0 | | | |
| T21_I3_TD | 76 | 0.44 | -0.53 | 1.16 | 0 | | | |
| T21_I9_TD | 76 | 0.20 | -0.99 | 1.36 | 0 | | | |
| T21_S3_TD | 76 | 0.55 | -0.11 | 0.87 | 0 | | | |
| T21_S9_TD | 75 | 0.51 | -0.52 | 1.15 | 1 | -1.17 | -1.17 | -1.17 |
