## Supplementary material for "Early Pointwise Sensitivity Fluctuation Predicts Glaucoma Progression": file:///Users/yuriribeiro/Downloads/Table%205.pdf

**Table 5.** Comparison of the run-in medians rate of VF sensitivity change (dB) of total deviation (TD) from each 24-2 visual field location, considering progressive and non-progressive events in the glaucoma suspect group.

| VF location | Non-progressive events | | | | Progressive events | | | | P-value* |
|---|---|---|---|---|---|---|---|---|---|
| | N (eyes) | Median | P25 | P75 | N (eyes) | Median | P25 | P75 | |
| N3_I3_TD | 363 | 0.16 | -0.40 | 0.70 | 1 | 0.01 | 0.01 | 0.01 | 0.8790 |
| N3_I9_TD | 363 | 0.11 | -0.46 | 0.62 | 1 | -1.10 | -1.10 | -1.10 | 0.1327 |
| N3_I15_TD | 359 | 0.22 | -0.46 | 0.76 | 5 | 0.48 | -0.70 | 1.43 | 0.5098 |
| N3_I21_TD | 355 | 0.10 | -0.65 | 0.82 | 9 | -0.23 | -4.05 | 0.95 | 0.3123 |
| N3_S3_TD | 362 | 0.13 | -0.42 | 0.68 | 2 | 0.26 | 0.10 | 0.42 | 0.8057 |
| N3_S9_TD | 360 | 0.10 | -0.41 | 0.63 | 4 | 0.30 | -0.17 | 0.46 | 0.7798 |
| N3_S15_TD | 360 | 0.08 | -0.55 | 0.68 | 4 | 0.12 | -0.80 | 0.82 | 0.9905 |
| N3_S21_TD | 348 | 0.26 | -0.53 | 1.24 | 16 | -1.01 | -2.26 | 0.50 | 0.0058 |
| N9_I3_TD | 360 | 0.06 | -0.32 | 0.59 | 4 | -0.22 | -0.88 | 0.21 | 0.2485 |
| N9_I9_TD | 361 | 0.01 | -0.39 | 0.53 | 3 | 0.00 | -0.05 | 0.74 | 0.6714 |
| N9_I15_TD | 359 | 0.10 | -0.32 | 0.70 | 5 | -0.60 | -6.22 | 2.97 | 0.5071 |
| N9_I21_TD | 357 | 0.16 | -0.67 | 0.97 | 7 | -2.08 | -3.94 | 1.88 | 0.2608 |
| N9_S3_TD | 361 | 0.15 | -0.36 | 0.55 | 3 | -0.74 | -1.54 | 0.75 | 0.3160 |
| N9_S9_TD | 363 | 0.10 | -0.40 | 0.56 | 1 | -0.37 | -0.37 | -0.37 | 0.3970 |
| N9_S15_TD | 362 | 0.18 | -0.43 | 0.71 | 2 | -1.28 | -2.48 | -0.08 | 0.1269 |
| N9_S21_TD | 352 | 0.14 | -0.67 | 1.04 | 12 | -1.62 | -4.73 | -0.32 | 0.0002 |
| N15_I3_TD | 360 | 0.06 | -0.51 | 0.69 | 4 | 0.49 | -1.57 | 3.11 | 0.7872 |
| N15_I9_TD | 359 | 0.01 | -0.50 | 0.62 | 5 | 1.07 | -2.94 | 1.33 | 0.5462 |
| N15_I15_TD | 353 | 0.09 | -0.60 | 0.77 | 11 | -0.88 | -3.50 | 0.80 | 0.0920 |
| N15_S3_TD | 360 | 0.09 | -0.56 | 0.67 | 4 | -0.56 | -5.96 | 1.17 | 0.3703 |
| N15_S9_TD | 361 | 0.04 | -0.50 | 0.70 | 3 | -0.29 | -0.65 | 1.05 | 0.8213 |
| N15_S15_TD | 355 | 0.18 | -0.54 | 0.97 | 9 | -1.24 | -2.26 | 0.79 | 0.0799 |
| N21_I3_TD | 355 | 0.10 | -0.46 | 0.84 | 9 | -0.27 | -3.56 | 0.98 | 0.4244 |
| N21_I9_TD | 357 | 0.13 | -0.71 | 0.85 | 7 | -2.79 | -5.79 | 3.89 | 0.0952 |
| N21_S3_TD | 357 | 0.17 | -0.46 | 0.84 | 7 | 0.33 | -1.20 | 1.43 | 0.8221 |
| N21_S9_TD | 357 | 0.20 | -0.76 | 1.12 | 7 | -1.09 | -3.26 | 1.25 | 0.1146 |
| N27_I3_TD | 351 | 0.13 | -0.73 | 0.92 | 13 | -0.01 | -2.55 | 0.24 | 0.0553 |
| N27_S3_TD | 351 | 0.20 | -0.51 | 1.19 | 13 | -1.19 | -4.24 | 1.48 | 0.2292 |
| T3_I3_TD | 363 | 0.06 | -0.40 | 0.58 | 1 | -0.94 | -0.94 | -0.94 | 0.1590 |
| T3_I9_TD | 362 | 0.01 | -0.50 | 0.56 | 2 | 0.86 | 0.48 | 1.24 | 0.1188 |
| T3_I15_TD | 359 | 0.02 | -0.54 | 0.66 | 5 | 0.27 | -2.43 | 1.22 | 0.9556 |
| T3_I21_TD | 362 | 0.23 | -0.55 | 0.87 | 2 | -0.84 | -1.23 | -0.45 | 0.1391 |
| T3_S3_TD | 362 | 0.02 | -0.43 | 0.74 | 2 | -0.11 | -0.22 | 0.00 | 0.6614 |
| T3_S9_TD | 360 | 0.05 | -0.52 | 0.59 | 4 | -0.79 | -1.38 | 0.02 | 0.0720 |
| T3_S15_TD | 358 | 0.16 | -0.51 | 0.86 | 6 | -0.78 | -1.94 | -0.35 | 0.0132 |
| T3_S21_TD | 348 | 0.03 | -0.72 | 1.00 | 16 | -0.90 | -2.89 | -0.46 | 0.0010 |
| T9_I3_TD | 361 | 0.07 | -0.41 | 0.72 | 3 | 0.82 | 0.00 | 1.97 | 0.1625 |
| T9_I9_TD | 360 | 0.00 | -0.49 | 0.52 | 4 | -0.23 | -0.93 | 0.33 | 0.4546 |
| T9_I15_TD | 358 | 0.09 | -0.44 | 0.68 | 6 | -0.13 | -2.16 | 0.28 | 0.2413 |
| T9_I21_TD | 357 | 0.13 | -0.54 | 0.77 | 7 | -0.60 | -2.71 | 0.58 | 0.0917 |
| T9_S3_TD | 361 | 0.03 | -0.49 | 0.69 | 3 | -0.68 | -4.88 | 0.00 | 0.0945 |
| T9_S9_TD | 360 | 0.13 | -0.41 | 0.65 | 4 | -0.18 | -1.37 | 0.25 | 0.2446 |
| T9_S15_TD | 359 | 0.03 | -0.59 | 0.80 | 5 | -1.17 | -3.43 | 0.44 | 0.0416 |
| T9_S21_TD | 346 | 0.16 | -0.81 | 1.05 | 18 | -0.95 | -2.18 | -0.02 | 0.0022 |

| | | | | | | | | | |
|---|---|---|---|---|---|---|---|---|---|
| T15_I9_TD | 360 | 0.05 | -0.55 | 0.82 | 4 | 0.69 | -0.88 | 1.47 | 0.6108 |
| T15_I15_TD | 361 | 0.10 | -0.53 | 0.76 | 3 | -1.03 | -2.52 | 1.25 | 0.3461 |
| T15_S9_TD | 358 | 0.18 | -0.56 | 0.84 | 6 | -1.86 | -3.78 | 0.01 | 0.0090 |
| T15_S15_TD | 357 | 0.10 | -0.57 | 1.01 | 7 | -1.32 | -3.16 | -0.67 | 0.0016 |
| T21_I3_TD | 358 | 0.11 | -0.49 | 0.71 | 6 | -0.87 | -4.42 | 0.44 | 0.0929 |
| T21_I9_TD | 357 | 0.16 | -0.56 | 0.82 | 7 | -0.73 | -2.76 | -0.04 | 0.0243 |
| T21_S3_TD | 357 | 0.10 | -0.58 | 0.74 | 7 | -0.65 | -1.82 | 0.85 | 0.2161 |
| T21_S9_TD | 361 | 0.25 | -0.69 | 0.86 | 3 | 0.31 | -0.41 | 0.55 | 0.9824 |

(*) Mann-Whitney test for the comparison of non-progressive and progressive events.
