## Supplementary material for "Early Pointwise Sensitivity Fluctuation Predicts Glaucoma Progression": file:///Users/yuriribeiro/Downloads/Table%204.pdf

**Table 4.** Comparison of the run-in medians rate of VF sensitivity change (dB) of total deviation (TD) from each 24-2 visual field location, considering progressive and non-progressive events in the glaucoma group.

| VF location | Non-progressive events | | | | Progressive events | | | | P-value* |
|---|---|---|---|---|---|---|---|---|---|
| | N (eyes) | Median | P25 | P75 | N (eyes) | Median | P25 | P75 | |
| N3_I3_TD | 183 | 0.07 | -0.38 | 0.64 | 18 | 0.30 | -1.18 | 1.91 | 0.4234 |
| N3_I9_TD | 174 | 0.10 | -0.36 | 0.83 | 27 | -0.21 | -1.10 | 0.24 | 0.0227 |
| N3_I15_TD | 170 | 0.08 | -0.50 | 0.83 | 31 | -0.61 | -1.83 | 0.37 | 0.0122 |
| N3_I21_TD | 166 | 0.23 | -0.60 | 0.92 | 35 | -0.80 | -2.02 | 0.92 | 0.0078 |
| N3_S3_TD | 181 | 0.00 | -0.43 | 0.70 | 20 | -0.61 | -2.29 | 0.85 | 0.1956 |
| N3_S9_TD | 165 | 0.00 | -0.53 | 0.70 | 36 | -0.64 | -2.70 | 0.72 | 0.0088 |
| N3_S15_TD | 167 | 0.18 | -0.57 | 1.14 | 34 | -0.92 | -1.91 | 0.41 | 0.0010 |
| N3_S21_TD | 156 | 0.09 | -1.21 | 1.23 | 45 | -0.21 | -1.79 | 1.03 | 0.2169 |
| N9_I3_TD | 179 | 0.07 | -0.33 | 0.72 | 22 | -0.27 | -1.86 | 0.92 | 0.0836 |
| N9_I9_TD | 179 | 0.00 | -0.45 | 0.58 | 22 | -1.00 | -2.08 | -0.20 | <.0001 |
| N9_I15_TD | 176 | 0.12 | -0.49 | 0.72 | 25 | -0.80 | -2.80 | 0.22 | 0.0008 |
| N9_I21_TD | 164 | 0.02 | -0.91 | 1.14 | 37 | -0.90 | -1.98 | 0.01 | 0.0013 |
| N9_S3_TD | 169 | 0.07 | -0.37 | 0.66 | 32 | -0.50 | -2.33 | 0.56 | 0.0158 |
| N9_S9_TD | 171 | 0.06 | -0.50 | 0.66 | 30 | -0.97 | -3.51 | 0.27 | 0.0010 |
| N9_S15_TD | 169 | 0.00 | -0.72 | 0.67 | 32 | -0.98 | -2.70 | 0.72 | 0.0340 |
| N9_S21_TD | 153 | 0.33 | -0.45 | 1.56 | 48 | -1.00 | -1.90 | 0.27 | <.0001 |
| N15_I3_TD | 178 | 0.13 | -0.52 | 0.77 | 23 | -0.28 | -3.15 | 0.59 | 0.0437 |
| N15_I9_TD | 174 | 0.04 | -0.55 | 0.58 | 27 | -0.88 | -2.12 | 0.31 | 0.0021 |
| N15_I15_TD | 170 | 0.17 | -0.56 | 0.85 | 31 | -0.65 | -2.13 | 0.75 | 0.0151 |
| N15_S3_TD | 166 | 0.10 | -0.32 | 0.74 | 35 | -0.90 | -3.57 | 0.17 | <.0001 |
| N15_S9_TD | 172 | 0.00 | -0.51 | 0.85 | 29 | -1.13 | -2.41 | 0.25 | <.0001 |
| N15_S15_TD | 155 | 0.00 | -0.74 | 0.86 | 46 | -0.97 | -2.16 | 0.50 | 0.0009 |
| N21_I3_TD | 166 | 0.20 | -0.72 | 0.93 | 35 | -0.27 | -1.34 | 0.56 | 0.0505 |
| N21_I9_TD | 167 | 0.14 | -0.85 | 0.95 | 34 | -0.73 | -3.65 | 0.93 | 0.0255 |
| N21_S3_TD | 166 | 0.00 | -0.73 | 0.88 | 35 | -0.86 | -1.83 | 0.16 | 0.0014 |
| N21_S9_TD | 165 | 0.18 | -0.67 | 1.30 | 36 | -0.62 | -1.71 | 0.59 | 0.0052 |
| N27_I3_TD | 162 | 0.27 | -0.59 | 1.69 | 39 | -1.39 | -3.08 | -0.13 | <.0001 |
| N27_S3_TD | 141 | 0.16 | -0.87 | 1.51 | 60 | -0.85 | -2.19 | 0.76 | 0.0008 |
| T3_I3_TD | 190 | 0.01 | -0.59 | 0.50 | 11 | -0.03 | -1.02 | 0.27 | 0.6756 |
| T3_I9_TD | 173 | 0.00 | -0.53 | 0.70 | 28 | -0.13 | -1.25 | 0.31 | 0.0627 |
| T3_I15_TD | 169 | -0.10 | -0.67 | 0.62 | 32 | -0.17 | -2.08 | 1.08 | 0.5192 |
| T3_I21_TD | 168 | 0.08 | -0.66 | 0.78 | 33 | -1.25 | -2.66 | 0.11 | <.0001 |
| T3_S3_TD | 176 | 0.14 | -0.36 | 0.76 | 25 | -0.52 | -3.97 | 0.53 | 0.0135 |
| T3_S9_TD | 161 | 0.00 | -0.54 | 0.60 | 40 | -0.43 | -2.76 | 0.25 | 0.0040 |
| T3_S15_TD | 167 | 0.20 | -0.62 | 1.08 | 34 | -0.77 | -3.65 | 0.09 | <.0001 |
| T3_S21_TD | 155 | 0.22 | -0.84 | 1.38 | 46 | -1.58 | -3.44 | 0.31 | <.0001 |
| T9_I3_TD | 190 | -0.00 | -0.63 | 0.49 | 11 | -0.05 | -1.02 | 0.48 | 0.6219 |
| T9_I9_TD | 175 | 0.05 | -0.47 | 0.66 | 26 | 0.04 | -1.28 | 0.66 | 0.5426 |
| T9_I15_TD | 174 | -0.02 | -0.61 | 0.55 | 27 | 0.07 | -0.87 | 1.01 | 0.4803 |
| T9_I21_TD | 169 | 0.24 | -0.63 | 0.86 | 32 | 0.17 | -1.56 | 0.75 | 0.1565 |
| T9_S3_TD | 169 | 0.09 | -0.51 | 0.69 | 32 | -0.71 | -2.11 | 0.78 | 0.0382 |
| T9_S9_TD | 168 | -0.01 | -0.47 | 0.77 | 33 | -0.24 | -1.45 | 0.58 | 0.0796 |
| T9_S15_TD | 166 | 0.10 | -0.48 | 0.62 | 35 | -0.44 | -1.73 | 0.47 | 0.0237 |
| T9_S21_TD | 150 | 0.01 | -0.76 | 1.26 | 51 | -0.91 | -3.05 | 0.32 | 0.0004 |

| | | | | | | | | | |
|---|---|---|---|---|---|---|---|---|---|
| T15_I9_TD | 177 | 0.18 | -0.43 | 0.88 | 24 | -0.09 | -1.70 | 0.77 | 0.1603 |
| T15_I15_TD | 176 | 0.08 | -0.73 | 0.89 | 25 | -0.47 | -1.65 | 0.47 | 0.1137 |
| T15_S9_TD | 164 | 0.00 | -0.70 | 0.80 | 37 | 0.22 | -1.44 | 1.37 | 0.6670 |
| T15_S15_TD | 168 | 0.24 | -0.67 | 1.06 | 33 | -1.40 | -2.98 | -0.56 | <.0001 |
| T21_I3_TD | 179 | 0.11 | -0.64 | 0.94 | 22 | -1.35 | -2.80 | -0.21 | <.0001 |
| T21_I9_TD | 180 | 0.12 | -0.67 | 1.01 | 21 | -0.37 | -1.04 | 0.25 | 0.0414 |
| T21_S3_TD | 177 | 0.22 | -0.57 | 0.85 | 24 | -0.86 | -1.76 | 0.41 | 0.0141 |
| T21_S9_TD | 176 | 0.21 | -0.83 | 0.95 | 25 | -1.03 | -1.99 | 0.39 | 0.0035 |

(*) Mann-Whitney test for the comparison of non-progressive and progressive events.
