## Supplementary material for "Early Pointwise Sensitivity Fluctuation Predicts Glaucoma Progression": file:///Users/yuriribeiro/Downloads/Table%203.pdf

**Table 3.** Comparison of the medians of run-in standard deviation of total deviation (TD) residuals (fluctuation, dB) from each 24-2 visual field test point location, by progression status in the control group.

| VF location | Non-progressive events | | | | Progressive events | | | |
|---|---|---|---|---|---|---|---|---|
| | N (eyes) | Median | P25 | P75 | N (eyes) | Median | P25 | P75 |
| N3_I3_TD | 76 | 0.93 | 0.55 | 1.39 | 0 | | | |
| N3_I9_TD | 76 | 1.00 | 0.70 | 1.39 | 0 | | | |
| N3_I15_TD | 76 | 1.04 | 0.70 | 1.65 | 0 | | | |
| N3_I21_TD | 75 | 1.29 | 0.75 | 1.94 | 1 | 6.89 | 6.89 | 6.89 |
| N3_S3_TD | 76 | 0.89 | 0.55 | 1.23 | 0 | | | |
| N3_S9_TD | 76 | 0.98 | 0.69 | 1.38 | 0 | | | |
| N3_S15_TD | 76 | 1.10 | 0.79 | 1.80 | 0 | | | |
| N3_S21_TD | 76 | 1.41 | 0.97 | 2.18 | 0 | | | |
| N9_I3_TD | 76 | 0.86 | 0.56 | 1.13 | 0 | | | |
| N9_I9_TD | 76 | 0.74 | 0.50 | 1.06 | 0 | | | |
| N9_I15_TD | 76 | 0.84 | 0.49 | 1.46 | 0 | | | |
| N9_I21_TD | 75 | 1.41 | 0.78 | 2.35 | 1 | 8.74 | 8.74 | 8.74 |
| N9_S3_TD | 76 | 0.87 | 0.49 | 1.27 | 0 | | | |
| N9_S9_TD | 76 | 0.85 | 0.48 | 1.29 | 0 | | | |
| N9_S15_TD | 76 | 0.97 | 0.57 | 1.47 | 0 | | | |
| N9_S21_TD | 74 | 1.71 | 0.99 | 2.69 | 2 | 2.02 | 0.49 | 3.55 |
| N15_I3_TD | 76 | 0.96 | 0.51 | 1.49 | 0 | | | |
| N15_I9_TD | 76 | 0.94 | 0.59 | 1.26 | 0 | | | |
| N15_I15_TD | 76 | 1.25 | 0.73 | 1.88 | 0 | | | |
| N15_S3_TD | 76 | 0.93 | 0.73 | 1.28 | 0 | | | |
| N15_S9_TD | 75 | 0.99 | 0.50 | 1.56 | 1 | 4.33 | 4.33 | 4.33 |
| N15_S15_TD | 76 | 1.39 | 0.77 | 2.05 | 0 | | | |
| N21_I3_TD | 76 | 1.19 | 0.73 | 1.93 | 0 | | | |
| N21_I9_TD | 76 | 1.31 | 0.76 | 2.07 | 0 | | | |
| N21_S3_TD | 76 | 1.09 | 0.79 | 1.68 | 0 | | | |
| N21_S9_TD | 75 | 1.40 | 0.82 | 2.40 | 1 | 6.75 | 6.75 | 6.75 |
| N27_I3_TD | 76 | 1.96 | 0.84 | 3.22 | 0 | | | |
| N27_S3_TD | 74 | 1.49 | 0.87 | 2.58 | 2 | 2.67 | 2.09 | 3.25 |
| T3_I3_TD | 76 | 0.97 | 0.51 | 1.36 | 0 | | | |
| T3_I9_TD | 76 | 0.91 | 0.54 | 1.31 | 0 | | | |
| T3_I15_TD | 76 | 0.93 | 0.69 | 1.27 | 0 | | | |
| T3_I21_TD | 75 | 1.21 | 0.81 | 1.68 | 1 | 1.40 | 1.40 | 1.40 |
| T3_S3_TD | 76 | 0.83 | 0.54 | 1.24 | 0 | | | |
| T3_S9_TD | 76 | 0.89 | 0.71 | 1.62 | 0 | | | |
| T3_S15_TD | 76 | 1.16 | 0.81 | 1.79 | 0 | | | |
| T3_S21_TD | 75 | 1.47 | 0.84 | 2.03 | 1 | 3.19 | 3.19 | 3.19 |
| T9_I3_TD | 76 | 0.99 | 0.79 | 1.50 | 0 | | | |
| T9_I9_TD | 76 | 0.83 | 0.43 | 1.23 | 0 | | | |
| T9_I15_TD | 76 | 0.98 | 0.57 | 1.43 | 0 | | | |
| T9_I21_TD | 76 | 1.27 | 0.83 | 1.72 | 0 | | | |
| T9_S3_TD | 76 | 0.99 | 0.58 | 1.43 | 0 | | | |
| T9_S9_TD | 76 | 0.74 | 0.42 | 1.12 | 0 | | | |
| T9_S15_TD | 76 | 0.96 | 0.56 | 1.43 | 0 | | | |
| T9_S21_TD | 73 | 1.60 | 1.03 | 2.67 | 3 | 4.53 | 0.95 | 9.00 |

| | | | | | | | | |
|---|---|---|---|---|---|---|---|---|
| T15_I9_TD | 76 | 0.98 | 0.65 | 1.70 | 0 | | | |
| T15_I15_TD | 76 | 1.29 | 0.74 | 1.89 | 0 | | | |
| T15_S9_TD | 76 | 1.09 | 0.77 | 1.80 | 0 | | | |
| T15_S15_TD | 76 | 1.46 | 0.86 | 2.53 | 0 | | | |
| T21_I3_TD | 76 | 1.24 | 0.88 | 1.73 | 0 | | | |
| T21_I9_TD | 76 | 1.10 | 0.71 | 1.75 | 0 | | | |
| T21_S3_TD | 76 | 1.40 | 0.84 | 1.94 | 0 | | | |
| T21_S9_TD | 75 | 1.30 | 0.83 | 2.05 | 1 | 2.62 | 2.62 | 2.62 |
