## Supplementary material for "Early Pointwise Sensitivity Fluctuation Predicts Glaucoma Progression": file:///Users/yuriribeiro/Downloads/Table%202.pdf

**Table 2.** Comparison of the medians of run-in standard deviation of total deviation (TD) residuals (fluctuation, dB) from each 24-2 visual field test point location, by progression status in the glaucoma suspect group.

| VF location | Non-progressive events | | | | Progressive events | | | | P-value* |
|---|---|---|---|---|---|---|---|---|---|
| | N (eyes) | Median | P25 | P75 | N (eyes) | Median | P25 | P75 | |
| N3_I3_TD | 363 | 0.87 | 0.57 | 1.19 | 1 | 0.82 | 0.82 | 0.82 | 0.8119 |
| N3_I9_TD | 362 | 0.84 | 0.49 | 1.20 | 1 | 0.32 | 0.32 | 0.32 | 0.1800 |
| N3_I15_TD | 359 | 0.92 | 0.58 | 1.36 | 5 | 1.05 | 0.58 | 1.72 | 0.6348 |
| N3_I21_TD | 355 | 1.22 | 0.83 | 1.76 | 9 | 2.43 | 0.88 | 3.20 | 0.0704 |
| N3_S3_TD | 362 | 0.86 | 0.50 | 1.24 | 2 | 1.28 | 1.19 | 1.36 | 0.1900 |
| N3_S9_TD | 360 | 0.89 | 0.55 | 1.29 | 4 | 0.84 | 0.33 | 1.55 | 0.7005 |
| N3_S15_TD | 360 | 1.14 | 0.76 | 1.70 | 4 | 1.50 | 1.00 | 5.82 | 0.2240 |
| N3_S21_TD | 348 | 1.37 | 0.87 | 2.26 | 16 | 2.12 | 1.10 | 3.92 | 0.0467 |
| N9_I3_TD | 360 | 0.77 | 0.48 | 1.14 | 4 | 0.44 | 0.11 | 1.01 | 0.1580 |
| N9_I9_TD | 361 | 0.76 | 0.44 | 0.99 | 3 | 0.44 | 0.00 | 1.89 | 0.6084 |
| N9_I15_TD | 359 | 0.94 | 0.56 | 1.32 | 5 | 1.60 | 0.92 | 2.37 | 0.0664 |
| N9_I21_TD | 357 | 1.23 | 0.74 | 1.94 | 7 | 2.14 | 1.41 | 5.23 | 0.0546 |
| N9_S3_TD | 361 | 0.80 | 0.52 | 1.21 | 3 | 0.42 | 0.23 | 1.24 | 0.2901 |
| N9_S9_TD | 363 | 0.82 | 0.49 | 1.11 | 1 | 0.91 | 0.91 | 0.91 | 0.7319 |
| N9_S15_TD | 362 | 1.05 | 0.63 | 1.55 | 2 | 2.76 | 2.65 | 2.88 | 0.0346 |
| N9_S21_TD | 352 | 1.55 | 0.94 | 2.46 | 12 | 2.27 | 1.58 | 3.37 | 0.0475 |
| N15_I3_TD | 358 | 0.86 | 0.53 | 1.22 | 4 | 0.98 | 0.74 | 5.15 | 0.3550 |
| N15_I9_TD | 358 | 0.89 | 0.53 | 1.29 | 5 | 3.03 | 1.54 | 4.94 | 0.0008 |
| N15_I15_TD | 353 | 1.14 | 0.82 | 1.71 | 11 | 2.35 | 1.29 | 3.91 | 0.0023 |
| N15_S3_TD | 360 | 0.86 | 0.55 | 1.28 | 4 | 1.75 | 0.55 | 2.69 | 0.2029 |
| N15_S9_TD | 361 | 0.94 | 0.57 | 1.35 | 3 | 2.59 | 1.48 | 6.63 | 0.0116 |
| N15_S15_TD | 355 | 1.35 | 0.83 | 2.04 | 9 | 2.36 | 1.25 | 4.26 | 0.0430 |
| N21_I3_TD | 355 | 0.96 | 0.65 | 1.56 | 9 | 2.37 | 1.34 | 4.61 | 0.0021 |
| N21_I9_TD | 357 | 1.29 | 0.84 | 2.01 | 7 | 4.71 | 0.40 | 7.13 | 0.0550 |
| N21_S3_TD | 357 | 0.97 | 0.60 | 1.51 | 7 | 1.71 | 1.35 | 6.41 | 0.0043 |
| N21_S9_TD | 357 | 1.29 | 0.81 | 2.00 | 7 | 2.03 | 1.32 | 3.10 | 0.0306 |
| N27_I3_TD | 351 | 1.44 | 0.83 | 2.69 | 13 | 1.76 | 1.19 | 3.07 | 0.3154 |
| N27_S3_TD | 351 | 1.48 | 0.83 | 2.42 | 13 | 2.53 | 1.22 | 3.19 | 0.0893 |
| T3_I3_TD | 363 | 0.82 | 0.51 | 1.22 | 1 | 5.55 | 5.55 | 5.55 | 0.0850 |
| T3_I9_TD | 362 | 0.86 | 0.54 | 1.23 | 2 | 0.60 | 0.24 | 0.96 | 0.3828 |
| T3_I15_TD | 359 | 0.95 | 0.59 | 1.37 | 5 | 1.30 | 0.68 | 2.41 | 0.2984 |
| T3_I21_TD | 362 | 1.21 | 0.75 | 1.71 | 2 | 0.81 | 0.34 | 1.28 | 0.3369 |
| T3_S3_TD | 361 | 0.86 | 0.53 | 1.22 | 2 | 0.95 | 0.00 | 1.90 | 0.9059 |
| T3_S9_TD | 360 | 0.92 | 0.56 | 1.38 | 4 | 1.53 | 0.51 | 1.79 | 0.3168 |
| T3_S15_TD | 358 | 1.03 | 0.64 | 1.67 | 6 | 2.07 | 1.11 | 3.54 | 0.0502 |
| T3_S21_TD | 348 | 1.53 | 1.01 | 2.33 | 16 | 2.24 | 1.65 | 3.11 | 0.0165 |
| T9_I3_TD | 361 | 0.94 | 0.65 | 1.38 | 3 | 1.00 | 0.00 | 2.76 | 0.9407 |
| T9_I9_TD | 359 | 0.76 | 0.48 | 1.07 | 4 | 1.02 | 0.35 | 2.18 | 0.7391 |
| T9_I15_TD | 358 | 0.87 | 0.53 | 1.25 | 6 | 0.78 | 0.00 | 1.96 | 0.7978 |
| T9_I21_TD | 357 | 1.20 | 0.77 | 1.65 | 7 | 0.98 | 0.78 | 2.70 | 0.7523 |
| T9_S3_TD | 361 | 0.95 | 0.56 | 1.37 | 3 | 1.85 | 0.00 | 4.83 | 0.4008 |
| T9_S9_TD | 360 | 0.88 | 0.54 | 1.29 | 4 | 1.00 | 0.48 | 1.41 | 0.8428 |
| T9_S15_TD | 358 | 1.06 | 0.69 | 1.51 | 5 | 0.88 | 0.75 | 1.33 | 0.7073 |
| T9_S21_TD | 346 | 1.73 | 1.07 | 2.62 | 18 | 2.11 | 1.58 | 4.22 | 0.0243 |

| | | | | | | | | | |
|---|---|---|---|---|---|---|---|---|---|
| T15_I9_TD | 360 | 1.00 | 0.59 | 1.41 | 4 | 2.08 | 0.42 | 5.33 | 0.3964 |
| T15_I15_TD | 361 | 1.03 | 0.64 | 1.54 | 3 | 2.46 | 0.52 | 7.07 | 0.2276 |
| T15_S9_TD | 358 | 1.09 | 0.75 | 1.67 | 6 | 2.08 | 0.91 | 2.75 | 0.0929 |
| T15_S15_TD | 357 | 1.29 | 0.85 | 1.91 | 7 | 1.53 | 1.06 | 1.93 | 0.6687 |
| T21_I3_TD | 358 | 1.11 | 0.75 | 1.62 | 6 | 1.94 | 1.49 | 5.11 | 0.0049 |
| T21_I9_TD | 357 | 1.14 | 0.78 | 1.65 | 7 | 1.71 | 1.28 | 1.99 | 0.0219 |
| T21_S3_TD | 357 | 1.10 | 0.71 | 1.66 | 7 | 1.57 | 0.79 | 1.79 | 0.2830 |
| T21_S9_TD | 361 | 1.12 | 0.75 | 1.81 | 3 | 2.20 | 1.44 | 3.53 | 0.0603 |

(*) Mann-Whitney test for the comparison of non-progressive and progressive events.
