## Supplementary material for "Early Pointwise Sensitivity Fluctuation Predicts Glaucoma Progression": file:///Users/yuriribeiro/Downloads/Table%201.pdf

**Table 1.** Comparison of the medians of run-in standard deviation of total deviation (TD) residuals (fluctuation, dB) from each 24-2 visual field test point location, by progression status in the glaucoma group.

| VF location | Non-progressive events | | | | Progressive events | | | | P-value* |
|---|---|---|---|---|---|---|---|---|---|
| | N (eyes) | Median | P25 | P75 | N (eyes) | Median | P25 | P75 | |
| N3_I3_TD | 183 | 0.89 | 0.57 | 1.24 | 18 | 1.64 | 0.96 | 7.18 | 0.0002 |
| N3_I9_TD | 174 | 0.86 | 0.49 | 1.41 | 27 | 1.28 | 0.74 | 2.41 | 0.0313 |
| N3_I15_TD | 170 | 1.14 | 0.60 | 1.68 | 31 | 1.68 | 0.92 | 2.53 | 0.0018 |
| N3_I21_TD | 166 | 1.36 | 0.90 | 2.07 | 35 | 1.82 | 1.18 | 2.58 | 0.0435 |
| N3_S3_TD | 181 | 0.86 | 0.55 | 1.37 | 20 | 1.37 | 0.68 | 2.32 | 0.0323 |
| N3_S9_TD | 165 | 1.04 | 0.58 | 1.67 | 36 | 1.69 | 1.05 | 3.67 | <.0001 |
| N3_S15_TD | 166 | 1.31 | 0.80 | 1.97 | 34 | 1.31 | 0.69 | 2.15 | 0.7660 |
| N3_S21_TD | 156 | 1.70 | 1.13 | 2.59 | 45 | 1.50 | 0.98 | 2.98 | 0.9594 |
| N9_I3_TD | 179 | 0.84 | 0.52 | 1.27 | 22 | 1.83 | 0.91 | 4.22 | <.0001 |
| N9_I9_TD | 179 | 0.82 | 0.51 | 1.18 | 22 | 1.28 | 0.83 | 2.21 | 0.0021 |
| N9_I15_TD | 176 | 1.11 | 0.71 | 1.78 | 25 | 1.67 | 0.80 | 2.96 | 0.0279 |
| N9_I21_TD | 164 | 1.64 | 0.98 | 2.52 | 37 | 1.64 | 1.10 | 3.57 | 0.3634 |
| N9_S3_TD | 169 | 0.90 | 0.49 | 1.30 | 32 | 1.65 | 0.95 | 4.20 | <.0001 |
| N9_S9_TD | 171 | 0.86 | 0.54 | 1.40 | 30 | 1.05 | 0.76 | 4.45 | 0.0124 |
| N9_S15_TD | 168 | 1.01 | 0.65 | 1.69 | 32 | 2.22 | 1.14 | 5.42 | <.0001 |
| N9_S21_TD | 153 | 1.72 | 0.94 | 2.80 | 48 | 1.79 | 1.01 | 4.20 | 0.2692 |
| N15_I3_TD | 178 | 1.04 | 0.70 | 1.54 | 23 | 1.75 | 0.82 | 3.20 | 0.0043 |
| N15_I9_TD | 174 | 0.99 | 0.67 | 1.48 | 27 | 1.65 | 0.94 | 3.24 | 0.0014 |
| N15_I15_TD | 170 | 1.39 | 0.81 | 1.99 | 31 | 1.98 | 1.29 | 2.87 | 0.0035 |
| N15_S3_TD | 166 | 0.97 | 0.57 | 1.65 | 35 | 1.72 | 0.96 | 3.95 | <.0001 |
| N15_S9_TD | 172 | 1.06 | 0.74 | 1.71 | 29 | 1.21 | 0.79 | 4.18 | 0.0455 |
| N15_S15_TD | 155 | 1.57 | 0.99 | 2.45 | 46 | 2.09 | 1.02 | 3.88 | 0.0719 |
| N21_I3_TD | 166 | 1.18 | 0.69 | 1.87 | 35 | 1.83 | 1.12 | 2.99 | 0.0042 |
| N21_I9_TD | 166 | 1.37 | 0.85 | 2.47 | 34 | 2.39 | 1.48 | 3.68 | 0.0002 |
| N21_S3_TD | 165 | 1.36 | 0.85 | 2.25 | 35 | 2.12 | 1.29 | 4.29 | 0.0008 |
| N21_S9_TD | 165 | 1.63 | 0.80 | 2.48 | 36 | 2.69 | 1.96 | 3.81 | 0.0004 |
| N27_I3_TD | 162 | 1.84 | 0.85 | 3.09 | 39 | 2.46 | 1.48 | 4.09 | 0.0380 |
| N27_S3_TD | 141 | 1.88 | 1.00 | 3.33 | 60 | 2.09 | 1.29 | 3.85 | 0.1901 |
| T3_I3_TD | 190 | 0.86 | 0.55 | 1.36 | 11 | 0.97 | 0.90 | 1.48 | 0.2709 |
| T3_I9_TD | 173 | 0.88 | 0.49 | 1.25 | 28 | 1.16 | 0.58 | 2.13 | 0.0871 |
| T3_I15_TD | 169 | 0.98 | 0.66 | 1.63 | 32 | 1.81 | 1.12 | 3.22 | <.0001 |
| T3_I21_TD | 168 | 1.26 | 0.84 | 1.82 | 33 | 1.50 | 1.05 | 3.45 | 0.0304 |
| T3_S3_TD | 176 | 0.95 | 0.56 | 1.32 | 25 | 1.50 | 0.79 | 5.61 | 0.0042 |
| T3_S9_TD | 161 | 1.09 | 0.56 | 1.63 | 40 | 1.52 | 0.93 | 2.21 | 0.0024 |
| T3_S15_TD | 166 | 1.27 | 0.73 | 1.91 | 34 | 1.51 | 0.99 | 3.34 | 0.0371 |
| T3_S21_TD | 155 | 1.62 | 0.86 | 2.93 | 46 | 2.35 | 1.64 | 3.73 | 0.0003 |
| T9_I3_TD | 190 | 0.97 | 0.52 | 1.51 | 11 | 1.50 | 0.97 | 1.92 | 0.0191 |
| T9_I9_TD | 175 | 0.95 | 0.57 | 1.24 | 26 | 1.03 | 0.76 | 4.01 | 0.1123 |
| T9_I15_TD | 174 | 0.97 | 0.55 | 1.48 | 27 | 1.91 | 1.22 | 3.56 | <.0001 |
| T9_I21_TD | 169 | 1.30 | 0.84 | 1.95 | 32 | 1.66 | 0.91 | 3.83 | 0.1221 |
| T9_S3_TD | 169 | 1.20 | 0.70 | 1.63 | 32 | 1.92 | 1.10 | 4.21 | <.0001 |
| T9_S9_TD | 168 | 1.01 | 0.64 | 1.51 | 33 | 1.94 | 0.98 | 4.31 | 0.0002 |
| T9_S15_TD | 166 | 1.23 | 0.75 | 1.94 | 35 | 1.89 | 1.31 | 4.43 | <.0001 |
| T9_S21_TD | 150 | 1.91 | 1.23 | 2.79 | 51 | 2.01 | 1.32 | 3.62 | 0.5297 |

| | | | | | | | | | |
|---|---|---|---|---|---|---|---|---|---|
| T15_I9_TD | 177 | 1.22 | 0.83 | 1.70 | 24 | 1.68 | 0.89 | 4.56 | 0.0135 |
| T15_I15_TD | 176 | 1.14 | 0.82 | 1.61 | 25 | 2.13 | 0.83 | 4.36 | 0.0051 |
| T15_S9_TD | 163 | 1.28 | 0.81 | 1.96 | 37 | 2.06 | 0.98 | 3.21 | 0.0055 |
| T15_S15_TD | 168 | 1.62 | 0.96 | 2.37 | 33 | 1.95 | 1.30 | 3.35 | 0.0464 |
| T21_I3_TD | 179 | 1.17 | 0.74 | 1.79 | 22 | 1.75 | 1.27 | 4.15 | 0.0012 |
| T21_I9_TD | 180 | 1.32 | 0.71 | 1.88 | 21 | 1.92 | 1.40 | 5.80 | 0.0014 |
| T21_S3_TD | 177 | 1.29 | 0.80 | 1.90 | 24 | 1.70 | 0.94 | 2.97 | 0.0613 |
| T21_S9_TD | 176 | 1.23 | 0.84 | 1.97 | 25 | 2.33 | 1.66 | 3.50 | <.0001 |

(*) Mann-Whitney test for the comparison of non-progressive and progressive events.
